# Multi-Cancer PRS Constellation Model for Cancer Risk Prediction

**DOI:** 10.1101/2024.10.17.24315686

**Authors:** Núria Moragas, Anna Díez-Villanueva, Ferran Moratalla-Navarro, Pablo Fernández-Navarro, Beatriz Pérez-Gómez, María Morales Suárez-Varela, Ana Molina-Barceló, Gemma Castaño-Vinyals, Blanca Rius-Sansalvador, Lois Riobó-Mayo, Rocío Olmedo-Requena, José-Juan Jiménez-Moleón, Rafael Marcos-Gragera, Marcela Guevara, Guillermo Fernandez-Tardon, Pilar Amiano Exezarreta, José M. Huerta, Tania Fernández-Villa, Antonio José Molina de la Torre, Vicente Martín-Sánchez, Inés Gómez-Acebo, Trinidad Dierssen, Juan Alguacil, Elisabet Guinó, Manolis Kogevinas, Marina Pollán, Mireia Obón-Santacana, Victor Moreno

## Abstract

Cancer development is influenced by genetic factors and modifiable exposures. GWAS has identified genetic variants and developed of prediction models through Polygenic Risk Scores (PRS), but PRS alone has limitations for estimating cancer risk.

This study assesses a novel PRS constellation approach that integrates Polygenic Risk Scores (PRS) from both lifestyle and genetic traits to enhance prediction models for colorectal, breast, and prostate cancers. The approach was developed using the UK Biobank dataset and validated in the independent GenRisk cohort.

The model, incorporating sex and age, achieved AUCs of 0.74 for CRC, 0.65 for BC, and 0.75 for PC in the UK Biobank. Including tumor-related PRSs improved PC prediction but had limited impact on CRC and BC. Age and sex inclusion boosted CRC and PC model accuracy. However, GenRisk validation showed reduced AUCs and limited utility of lifestyle PRSs, with CRC and BC models achieving 0.62 and PC 0.56.

Integrating lifestyle-related characteristics into PRS does not significantly enhance cancer-specific PRS prediction. However, PRSs for these traits show independent predictive power, highlighting the importance of considering lifestyle in cancer risk and the need for precision medicine to improve early detection.

## Inroduction

Cancer continues to exert a profound impact on global health, standing as one of the leading causes of morbidity and mortality worldwide. In 2020 alone, the disease wrought staggering numbers, with an estimated 19.3 million new cases and claiming nearly 10 million lives. Colorectal, breast, and prostate cancers are the most frequent among the European population, accounting for a total of 41.1% of all cases, representing a total of 5.6 million cases(1). Therefore, studying these types of cancer is crucial for developing effective prevention and treatment strategies, as well as for conducting cancer risk assessment to guide targeted screening efforts, which play a crucial role in early detection and prevention.

When considering screening options for colorectal cancer (CRC), such as colonoscopies and fecal immunochemical testing (FIT), it is important to weigh both cost-effectiveness and accuracy. While more economical FIT has its merits, the more expensive and invasive colonoscopy offers higher precision in detecting CRC. Similarly, in the case of breast cancer (BC) screening, mammography is the primary tool, but it is known to have a significant false-negative rate, particularly in women aged 40-49. For prostate cancer (PC), prostate-specific antigen (PSA) tests face challenges such as false negatives and concerns about overdiagnosis. These common cancers would benefit from improved screening strategies that consider both genetic and lifestyle factors, allowing for a more personalized and comprehensive approach to cancer risk management and prevention.

Cancer development is a multifactorial complex process influenced by various factors, including heritable genetic factors and modifiable exposures (e.g., environment, unhealthy dietary habits, stress, smoking, sedentary lifestyle, as well as social factors such as age, sex and ancestry)(2). Over the past few decades, genome-wide association studies (GWAS) have made remarkable progress in identifying numerous genetic risk variants associated with different types of cancer, as well as the development of prediction models through the calculation of Polygenic Risk Scores (PRS)(3–5). While these findings have expanded the knowledge of the genetic factors contributing to cancer susceptibility, it is essential to recognize and examine the limitations of relying solely on PRS from GWAS for estimating cancer risk.

Multiple studies have consistently shown that unhealthy lifestyles are the main contributors to the high burden of CRC, BC and PC(6–8). However, the impact of any factor alone is limited, and a comprehensive approach is required.

In this study, we present a novel methodology for assessing the contribution of various factors to cancer risk. Our approach involves integrating PRS specific to the cancers under investigation (CRC, BC, and PC) with genetically predicted lifestyle variables, thereby constructing a comprehensive PRS constellation model. The primary objective is to enhance the accuracy and personalization of risk assessment for each disease, by employing PRS constellations in the context of these cancers, as well as to explore the potential benefits and limitations of its use. Our findings shed light on the potential applications of PRS in the context of these cancers and highlight the need for further research in this area.

## Methods

The study followed a four-step process. First, GWAS studies were reviewed for CRC, BC, and PC, as well as single nucleotide polymorphisms (SNPs) linked to lifestyle variables considered risk factors for each cancer. Second, polygenic risk scores (PRS) with these SNPs were computed in the UK Biobank data, generating a genetic prediction for each cancer type and lifestyle variable considered. Third, Bayesian networks were fitted to model the risk of each cancer with all the PRSs, that we call PRS constellation. These network models allowed exploring the association of genetically predicted variables with cancer risk and among them, as well as the improvement of predictive accuracy. Finally, the model’s robustness was validated using an independent dataset (GenRisk, https://cancer.genrisk.org).

### Study populations

The UK Biobank (UKB - https://www.ukbiobank.ac.uk/) is a large, ongoing cohort study with genetic, clinical, and lifestyle data from 502,463 UK participants (2006-2010). The study design and methods have been previously described(9). For this study, we selected incident CRC, BC (only in woman), and PC cases. Cases and controls were defined as “white British” individuals of European ancestry. Prevalent cases, BC in men, and participants with poor genotyping data or study discontinuation were excluded. A random selection of 10 control subjects free of cancer was performed for each cancer outcome. The study comprised 5288 CRC cases, 7765 BC cases, and 8733 PC cases, and 52880, 77650, and 87330 controls respectively for each type of cancer. This research was conducted using the UK Biobank Resource (UKBv3) under Application 69033.

GenRisk is a sub-project of the MCC-Spain, a multicentre case-control study conducted between 2008 and 2013 in 12 provinces of Spain(10). The study included among other, cases of CRC, BC, PC, and share a common control population. For this study we selected a total of 6,387 subjects with genotype data, comprising 1,431 cases with CRC, 1,179 with BC, 913 with PC and 2,864 controls(11).

### Risk factors and Polygenic Risk Score (PRS) selection

Selection of risk factors for CRC, BC and PC was based on bibliography(6–8). A total of 63 traits of interest were considered for exploration, that were divided into 10 categories: alcohol consumption, smoking consumption, anthropometric body measurements, physical activity, dietary habits, supplement medicaments intake, medical history, cardiac function, women’s/men health, and biomarkers.

The PRS for CRC, BC, PC, and for each risk factor variable were developed using lists of SNPs derived from pre-existing PRSs proposed in other studies. These PRSs were obtained from the *PGS catalog* database(12) (https://www.pgscatalog.org/) and PubMed publications(13,14). The selection was based on the most recent studies and with the highest percentage of participants with European ancestry. SNPs on the sex chromosomes were excluded from PRS. Supplementary Table 1 provides a comprehensive overview of the variables used to calculate the PRSs. The UKB was the unique source for the risk factor PRS, and contributed to all the cancer-specific PRSs.

### Genotype data and imputation

The UKB and GenRisk data followed different procedures in terms of genotyping and imputation. UKB v3 imputed genotype data from 488,000 participants were obtained from their website, the genotyping and imputation protocol has been described elsewhere(9). Briefly, samples were genotyped using two arrays the UK BiLEVE Axiom Array (∼50000 participants) and UK Biobank Axiom Array (∼450000 participants) by Affymetrix. As a result, of the combination of the two arrays, 805426 markers were obtained in the + stand and in GRCh37 coordinates. Genotyped data was imputed using the Haplotype Reference Consortium (HRC) and UK10K reference panels, ∼96 million variants were obtained which are stored in BGEN v1.2 format (.bgen, .sample, .bgi). SNPs associated with the risk variables were extracted using the bgnix (https://enkre.net/cgi-bin/code/bgen/doc/trunk/doc/wiki/bgenix.md) program, and transformed into PLINK2 format (pgen, .pvar, .psam).

The GenRisk study was genotyped using two platforms. Infinium Oncoarray-500k was used for CRC cases an controls that were included in the CORECT(15) project and PC cases and controls included in the PRACTICAL(16), both part of the GAME-ON project(17), and Illumina Global Screening Array-24 v3.0 for the remaining cases and controls. A description of the sample types associated with each project is available in Supplementary Table 2. Quality control (QC) measures included sample removal based on call rate, sex concordance, heterozygosity, and duplicate handling. SNPs were filtered based on chromosomal location, allele validity, multi-locus nature, minor allele frequency, and call rate. Individual imputations were performed using the HRC panel for each of the subsets and subsequently merged selecting SNPs with good imputation quality (r2>0.4) in all platforms. The extraction of SNPs associated with risk variables and data manipulation were accomplished using *bcftools (*https://samtools.github.io/bcftools/bcftools.html)(18), and *PLINK2*(19).

### Construction and Assessment of PRS

The risk SNPs extracted from the imputed data from the UKB and GenRisk studies were subjected to rigorous QC measures prior to PRS calculation. The QC process encompassed the following steps: 1) Exclusion of multi-allelic, duplicated, and ambiguous SNPs with effective allele frequencies (EAF) ranging between 0.4 and 0.6. 2) Exclusion of variants with low imputation information (r2 < 0.4). 3) Exclusion of SNPs exhibiting a minor allele frequency (MAF) below the threshold of 0.005. 4) Exclusion of SNPs with high linkage disequilibrium (LD), using a correlation threshold (R2) of 0.3. When multiple SNPs were in LD, one was randomly selected and those with R2>0.3 were excluded, and 5) The Hardy-Weinberg Equilibrium (HWE) filter was applied to exclude variants exhibiting deviations from equilibrium, showing a statistical significance (p<1xE^-6^) in the HWE test.

Weighted PRSs were calculated using PLINK2 --score command, which calculates the lineal risk scores for each individual (*j* in the formula below) as a weighted sum of the *i* trait associated SNPs divided by the number of non-missing alleles, following the protocol defined for Collister *et al.* 2022(20), and using the following formula:

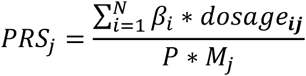

Where:

- N - The total number of SNPs.
- *β*_*i*_ - The SNPs weight
- *dosage*_*ij*_ - The allelic dosage for the effect allele in SNP ***i*** for individual ***j***.
- *P* - The ploidy of the individuals (*P* = 2 in humans).
- *M*_*j*_ - The number of non-missing variants observed for individual ***j***.

The PRS calculation used weights obtained from the SNPs effect sizes in the GWAS study that defined the PRS (Supplementary Table 1).

### Statistical analysis

All statistical analysis were performed using R software (version 4.2.0) and PLINK software (version 2.0). For simple comparisons between cases and controls, the chi-square test and Mann-Whitney U test were used. Pearson correlation was used to assess the association among quantitative variables.

### PRS Constellation prediction model building

A Bayesian network (BN) was constructed based on PRS calculated for each cancer type. The networks included as candidate nodes all the PRSs and were directed to predict the actual case/control status for each cancer. The structure learning of the BN was performed using the Hill-Climbing (hc) algorithm, which is a score-based learning approach. The *hc()* function from the *bnlearn* package in R was employed for this purpose. The learning process was guided by the Akaike Information Criterion (AIC) score, which was used to evaluate the goodness of fit of different network structures. The UKB was utilized for the structure learning process, enabling the identification of the optimal BN structure. To perform this procedure, the UKB data was initially partitioned into two distinct subsets: a train subset and a test subset. Subsequently, 10 train/test datasets were generated from the train subset utilizing a resampling technique. Each of these 10 datasets was employed for constructing a BN. From the models generated, the PRS associated with variables deemed significant in 8 out of 10 models were identified. These selected variables were utilized in the formulation of the ultimate PRS constellation model. The BN graphical structure was constructed to directed acyclic graph (DAG) using the *viewer()* function from *bnviewer* package. The *ggvenn* package in R were used to create a Venn diagram of pleiotropic associations between cancer-related SNPs and trait-associated SNPs.

The predictive discrimination of the models was assessed with the area under the curve (AUC) of the receiver operating characteristic (ROC) curve in the UKB test subset and in the completely independent GenRisk study. The DeLong test was used to compare the AUCs. The *pROC* package in R were used to create the calibration curve and the ROC graph. Optimal cutoffs to maximize sensitivity and specificity in the UKB dataset were calculated and applied to the GenRisk study to calculate accuracy.

## Results

### General characteristics of study population

The baseline characteristics of the UKB participants in the PRS constellation model development, spanning CRC, BC, and PC cancers are summarized in Table 1. The CRC set (5288 cases, 52880 controls) had a median age of 62 for cases and 56 for controls, with CRC cases showing associations with higher tobacco and red meat consumption, lower vegetables, and fruit intake, and elevated diabetes and hypertension rates. The BC set (7765 cases, 77650 controls) showed that cases, compared to controls, tend to be older, smokers, and consume more red meat. The PC cohort (8733 cases, 87330 controls) comprised only men, with cases that had higher consumption of tobacco, more frequent diabetes, hypertension, and high cholesterol levels.

**Table 1.**
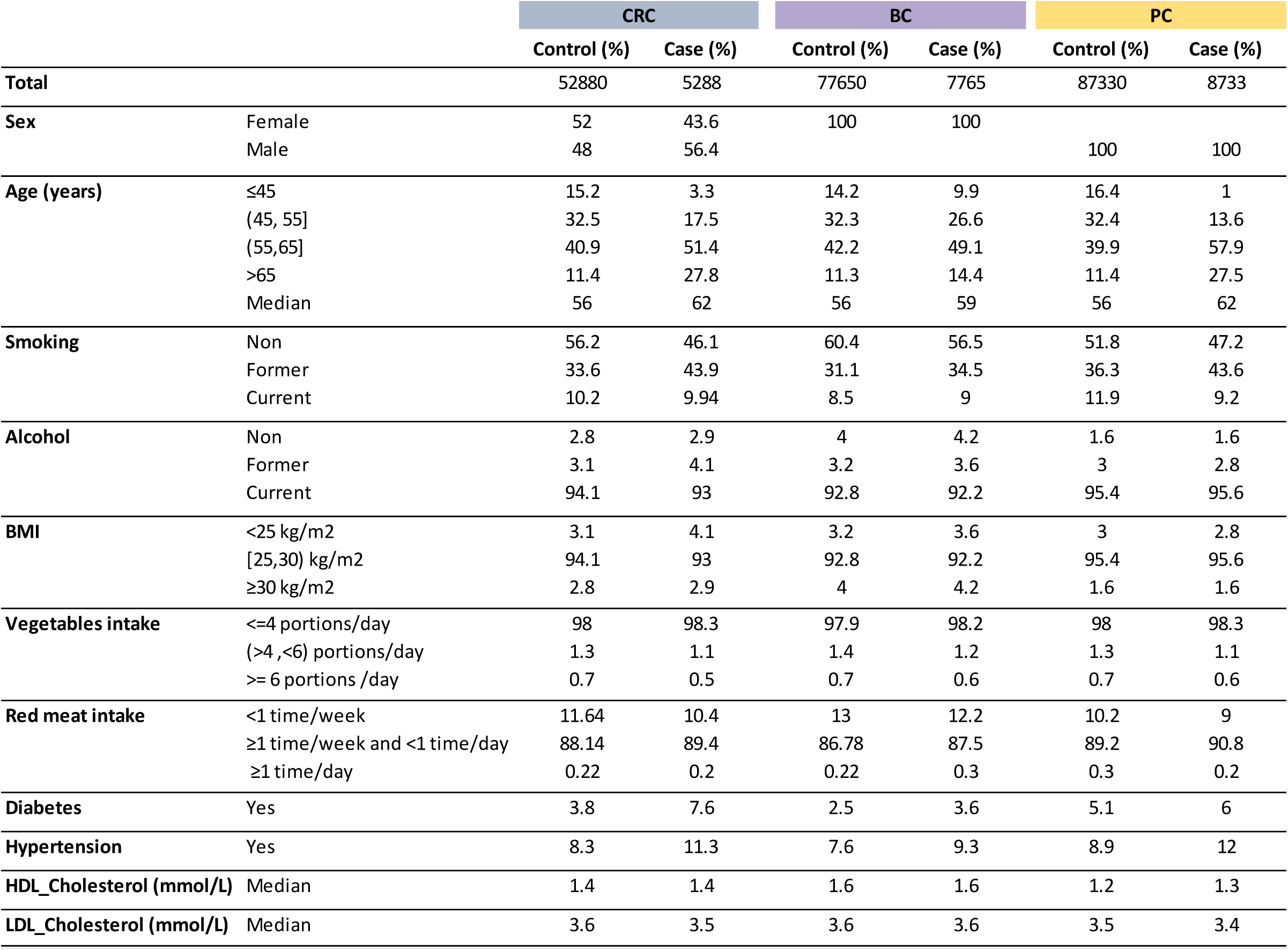
Characteristics of the UK Biobank study participants. CRC: Colorectal cancer; BC: Breast cancer; PC: Prostate cancer; BMI: Body Mass Index; Red meat intake includes Beef, Poultry and Lamb/mutton; Diabetes: diabetes diagnoses by doctor.

### Construction of PRS constellation model

A total of 63 weighted PRSs were created and evaluated for each cancer type and risk factor. Supplementary Figure 1 shows risk score distributions for CRC, BC, and PC, comparing cases and controls. The predictive power for the PRS evaluation of each trait is shown in Supplementary Table 3, demonstrating significant differences between case and control scores for all variables.

These PRSs were integrated into Bayesian network models, directed to predict the actual case status for each cancer type. The Bayesian network model was pruned from a fully connected model to identify variables influencing the target cancer and assigned coefficients to quantify their association strength. Age, and sex in the CRC model, were fixed in all models to ensure adjustment.

The constellation model for CRC was developed using a combination of 51 PRS, sex, and age variables, and the final model included 9 PRSs influencing CRC risk (Figure 1A). The mathematical formula was: CRC risk score = (0.0289 * Sex) + (0.0067 * Age at recruitment) + (0.0344 * PRS_CRC) – (0.0033 * PRS_Never_smoker) + (0.0047 * PRS_Standing_height) + (0.0042 * PRS_WHR) – (0.0036 * PRS_Aspirin) + (0.003 * PRS_Diabetes) + (0.006 * PRS_essential_Hypertension) + (0.004 * PRS_LDL_Cholesterol) + (0.004 * PRS_Pulse_rate_AR) – (0.0036 * PRS_Red_blood_cell_count). Figure 1B represents this PRS constellation model in a visualization of the relationships and dependencies among the variables, highlighting those influential in cases of CRC.

**Figure 1.**
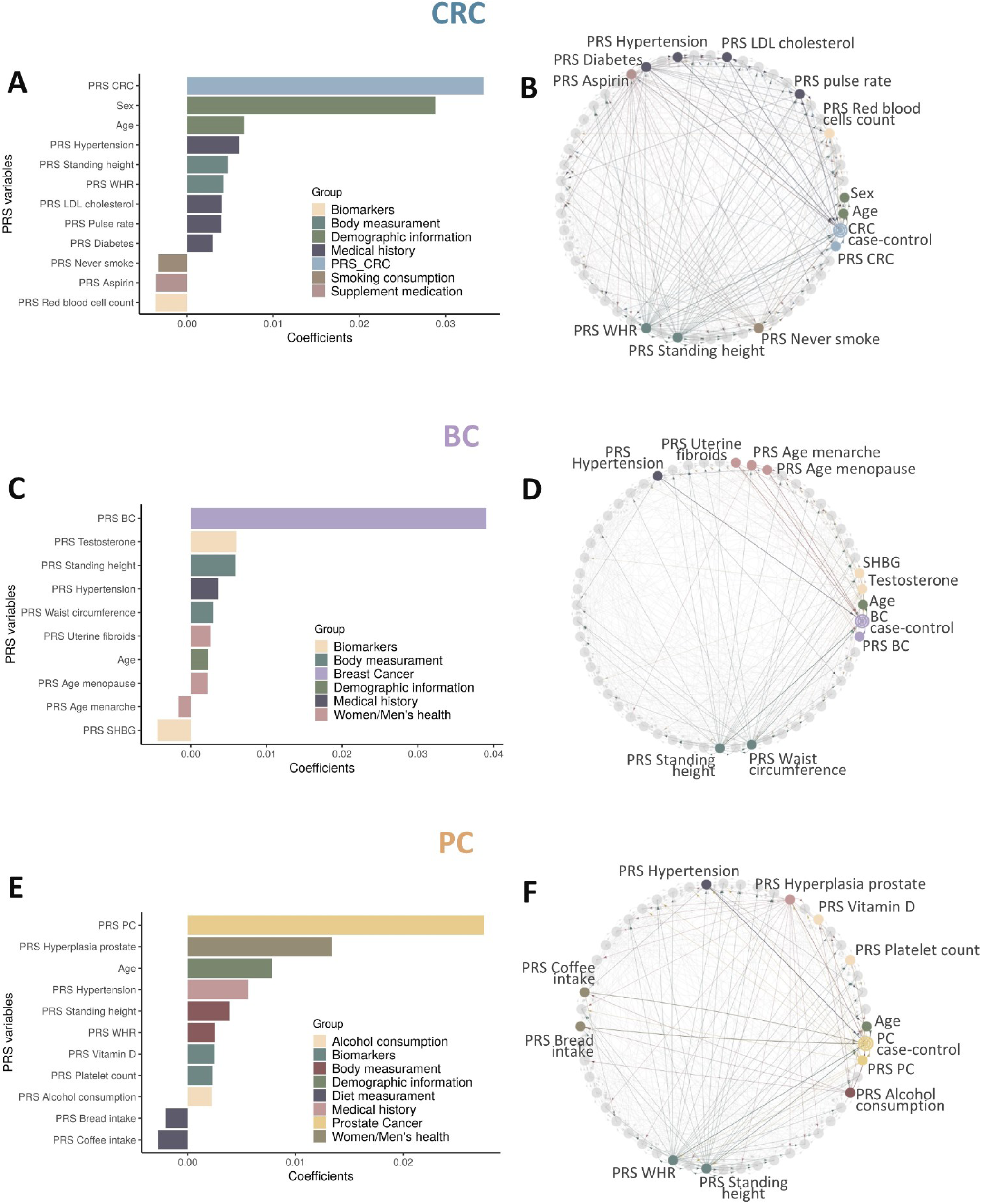
PRS Constellation Models for CRC, BC, and PC Risk Prediction. The figure displays the PRS constellation models and its coefficients for **A,B)** CRC, **C,D)** BC, and **E,F)** PC risk prediction. **A, C, and D)** Coefficients associated with each PRS variable in the Bayesian prediction models, sorted by their influence. Colours represent the group to which each variable belongs. **B, D, and F)** The models were constructed using the bnviewer package in R, which enables the visualization of Bayesian networks. The nodes in the figure represent a specific factor that contributes to the overall risk prediction. The arrows connecting the nodes depict the relationships and dependencies among the variables. The direction of the arrows indicates the direction of influence or causal relationships between the variables.

The BC PRS constellation model (Figures 1C and 1D) obtained using a combination of 54 PRS and age variable included 9 PRSs: BC risk score = (0.0023 * Age at recruitment) + (0.0391 * PRS_BC) + (0.0029 * PRS_Waist_circumference) + (0.0059 * PRS_Standing_height) + (0.0036 * PRS_essential_Hypertension) + (0.0026 * PRS_Uterine_fibroids) - (0.0016 * PRS_Age_menarche) + (0.0022 * PRS_Age_menopause) – (0.0044 * PRS_SHBG) + (0.006 * PRS_Testosterone).

The PC PRS constellation model (Figures 1E and 1F) was derived by incorporating a combination of 50 PRS variables along with the age variable, and included 10 PRSs: PC risk score = (0.0078 * Age at recruitment) + (0.0275 * PRS_PC) + (0.0022 * PRS_Alcohol_consumed) + (0.0039 * PRS_Standing_height) + (0.0025 * PRS_WHR) – (0.0018 * PRS_Bread_intake) - (0,0028 * PRS_Coffee_intake) + (0.0056 * PRS_essential_hypertension) + (0.0134 * PRS_hyperplasia_prostate) + (0.0025 * PRS_Vitamin_D_BM) + (0.0023 * PRS_Platelet_count_BM).

### Evaluation of cancer PRS constellation model using AUC analysis in the UKB test dataset

The three generated models underwent 10-fold cross-validation to assess their prediction power. ROC curves, illustrating sensitivity and specificity, were used alongside the AUC to evaluate the models’ discriminatory ability in predicting CRC, BC, and PC cancers. CRC model achieved an AUC of 0.74 (95%CI 0.72-0.76), with 60% sensitivity, 74% specificity, and 73% overall accuracy for a cutoff or 0.13. The BC model had an AUC of 0.65 (95%CI 0.63-0.67), yielding moderate sensitivity (56%), specificity (65%), and 64% overall accuracy for the 0.10 cutoff. The PC model obtained an AUC of 0.75 (95%CI 0.74-0.76), with 72% sensitivity, 66% specificity, and 66% overall accuracy for the 0.12 cutoff (Table 2, Supplementary Table 4 and Supplementary Figure 2).

**Table 2.** Predictive performance metrics of prediction models for CRC, BC, and PC in the UKB and GenRisk studies.

| PRS model | Included | Included | UKB |  |  | GenRisk |  |  |
| --- | --- | --- | --- | --- | --- | --- | --- | --- |
|  |  |  | AUC | 95% CI | p.value* | AUC | 95% CI | p.value* |
| CRC | sex & age | All | 0.74 | 0.72 - 0.76 | 6.2E-113 | 0.65 | 0.63 - 0.67 | 8.24E-39 |
|  |  | CRC | 0.74 | 0.72 - 0.76 | 1.2E-08 | 0.65 | 0.63 - 0.67 | 1.13E-38 |
|  |  | Traits | 0.70 | 0.69 - 0.72 | 5.2E-11 | 0.61 | 0.59 - 0.64 | 8.01E-23 |
|  | not included | All | 0.63 | 0.61 - 0.65 | 2.3E-36 | 0.62 | 0.59 - 0.64 | 9.05E-24 |
|  |  | CRC | 0.63 | 0.61 - 0.65 | 1.1E-35 | 0.62 | 0.60 - 0.64 | 4.83E-25 |
|  |  | Traits | 0.54 | 0.51 - 0.56 | 3.9E-04 | 0.50 | 0.48 - 0.53 | 0.37 |
| BC | age | All | 0.65 | 0.63 - 0.66 | 4.0E-63 | 0.56 | 0.54 - 0.58 | 1.6E-07 |
|  |  | BC | 0.65 | 0.63 - 0.67 | 2.4E-60 | 0.56 | 0.54 - 0.58 | 2.41E-07 |
|  |  | Traits | 0.57 | 0.55 - 0.59 | 1.6E-15 | 0.55 | 0.53 - 0.58 | 0.99 |
|  | not included | All | 0.64 | 0.62 - 0.65 | 3.1E-52 | 0.62 | 0.60 - 0.64 | 5.33E-25 |
|  |  | BC | 0.63 | 0.61 - 0.65 | 8.4E-50 | 0.62 | 0.60 - 0.64 | 5.21E-26 |
|  |  | Traits | 0.54 | 0.52 - 0.56 | 1.7E-06 | 0.51 | 0.49 - 0.54 | 0.11 |
| PC | age | All | 0.75 | 0.74 - 0.76 | 3.7E-197 | 0.50 | 0.47 - 0.53 | 0.53 |
|  |  | PC | 0.74 | 0.73 - 0.76 | 8.1E-187 | 0.50 | 0.46 - 0.53 | 0.45 |
|  |  | Traits | 0.73 | 0.72 - 0.75 | 4.9E-170 | 0.54 | 0.50 - 0.57 | 0.98 |
|  | not included | All | 0.60 | 0.59 - 0.62 | 7.2E-35 | 0.56 | 0.53 - 0.59 | 3.17E-04 |
|  |  | PC | 0.59 | 0.57 - 0.6 | 4.6E-25 | 0.58 | 0.55 - 0.62 | 7.88E-07 |
|  |  | Traits | 0.54 | 0.52 - 0.55 | 8.5E-06 | 0.53 | 0.49 - 0.56 | 0.93 |

### Impact on the accuracy of risk prediction when combining tumour related PRS traits with specific cancer PRS

While individual PRSs for traits demonstrated limited impact on overall risk (Supplementary Figure 3), a comprehensive evaluation was conducted to assess their combined influence. This evaluation compared models incorporating all PRSs (All-model), cancer-specific PRSs (Cancer-model), and trait-specific PRSs (Traits-model), both with and without the inclusion of sex and age. The findings revealed that the combined PRS traits did not significantly enhance the CRC or BC models when including or not age and sex. Nevertheless, a significant improvement was observed in the PC model when trait PRSs were combined with the cancer-specific PRS (Figure 2 and Table 2).

**Figure 2:**
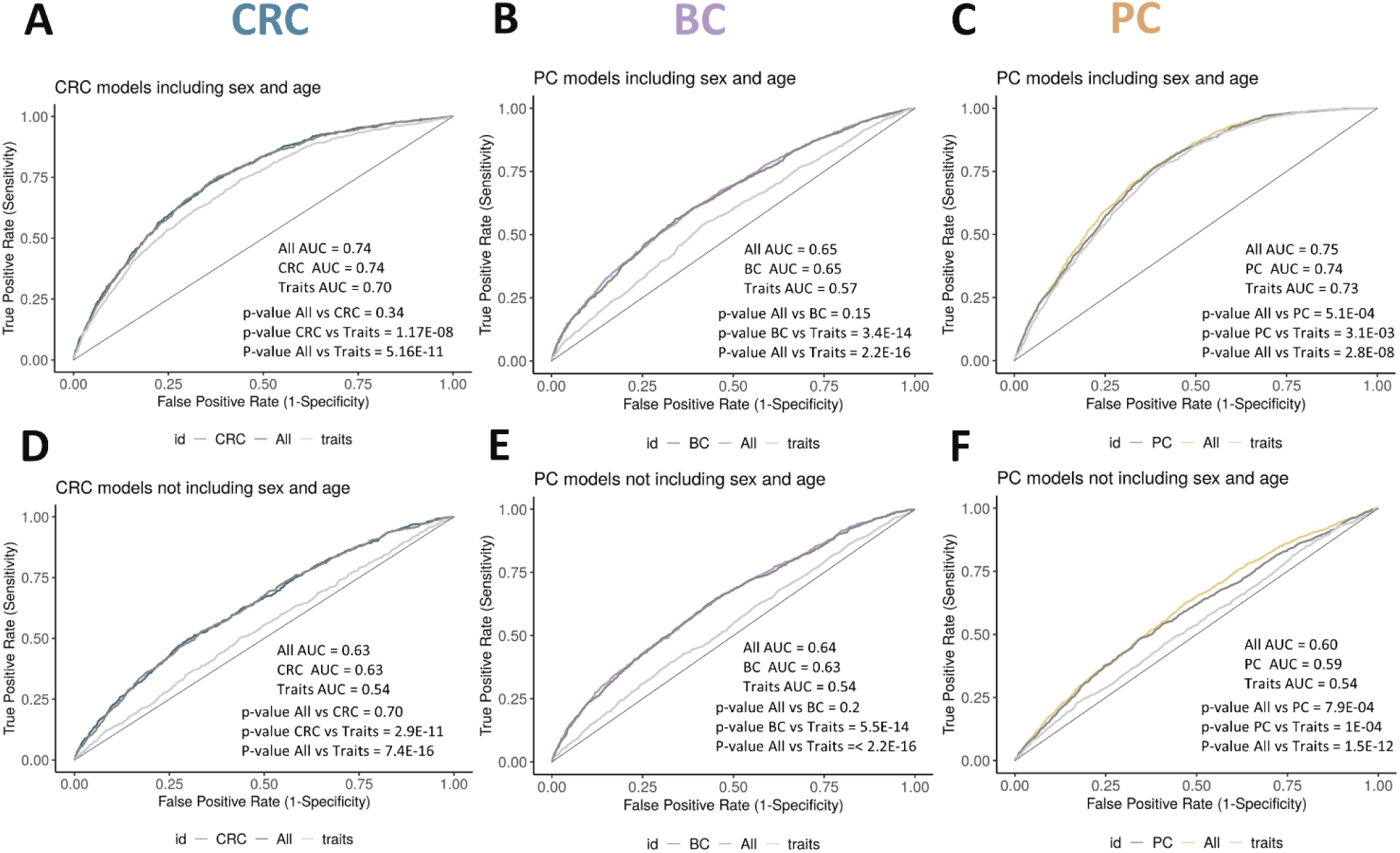
Comparative analysis of predictive power among models: PRS by Cancer (Cancer-model), PRS for case-related traits (Traits-model), and complete models (All-model). Evaluation includes ROC curves and AUC values for prediction models of **A)** Colorectal Cancer (CRC), **B)** Breast Cancer (BC), and **C)** Prostate Cancer (PC), including age and sex. Panels D-F represent ROC curves and AUC values for prediction models of **D)** CRC, **E)** BC, and **F)** PC, not including age and sex.

Remarkably, the traits-model demonstrated predictive capacity for all cancers, albeit to a lesser extent than the models incorporating cancer-specific PRSs. On the other hand, it should be noted that including age and sex variables in the model notably enhanced the AUC, especially in the CRC models, and age in the PC model (Figure 2 and Table 2).

Additionally, potential pleiotropic effects between cancer-related SNPs and trait-associated SNPs were analysed, indicating minimal overlap. Shared SNPs constituted a small fraction of the total SNP count (Supplementary Figure 4-6).

### Validation of PRS Constellation Models in the GenRisk study

The GenRisk study was utilized for the validation of the three risk prediction models, encompassing the comprehensive models (All-model), as well as an evaluation of the predictive potential of the specific PRS of each cancer (Cancer-model) and the PRSs associated with individual characteristics (Trait-model), both including and excluding age and sex.

In the external validation cohort of the CRC model including sex and age, we observed robust predictive accuracy for CRC, yielding an AUC value of 0.65 (95% CI 0.63-0.67) (Table 2). The model demonstrated a sensitivity of 70%, specificity of 50%, and an overall accuracy of 56%.. Additionally, it has been found that both the CRC-specific PRS (case-model) and the PRS associated with CRC-related variables (traits-model), in isolation, exhibited notable predictive capabilities for CRC, as evidenced by AUC values of 0.65 (95% CI 0.63-0.67) and 0.61 (95% CI 0.59-0.64), respectively. Thus, as observed in the UKB cohort, adding PRS traits to the CRC case-model did not improve its performance (Figure 3A, Table 2). In fact, the traits-model lacked predictive power when the variables age and sex were not included.

**Figure 3.**
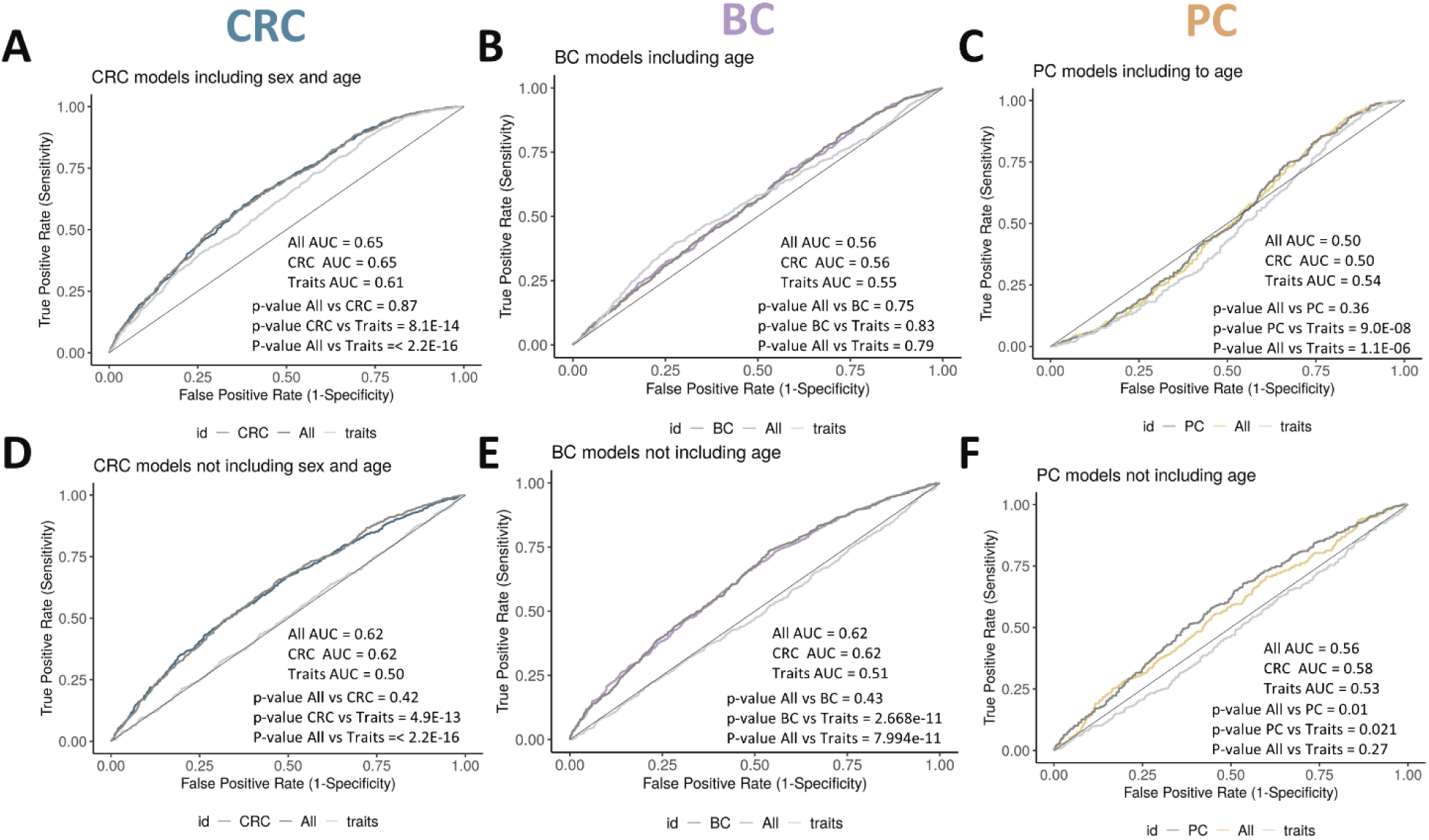
Discrimination ability of the PRS constellation models in the external validation cohort GenRisk. ROC curves of PRS constellations models: PRS by case (Case-model), PRS for case-related traits (Traits-model), and complete models (All-model). Evaluation includes ROC curves and AUC values for prediction models of **A)** Colorectal Cancer (CRC), **B)** Breast Cancer (BC), and **C)** Prostate Cancer (PC), including age and sex. Panels **D-F)** represent ROC curves and AUC values for prediction models of **D)** CRC, **E)** BC, and **F)** PC, not including age and sex.

The external validation of the BC All-model, including age, showed an AUC of 0.56 (95% CI 0.54-0.58) (Table 2), with a sensitivity of 52%, specificity of 54%, and overall accuracy of 53%. Similarly, the BC case-model, including age, demonstrated predictive capability with an AUC of 0.56 (95% CI 0.54-0.58) but the traits-model, even including age, did not exhibit significant predictive power, with an AUC of 0.55 (95% CI 0.53 - 0.58) (Figure 3B, Table 2). On the other hand, when age is excluded from the models, the AUC of both the all-model and the BC-model surprisingly improved to 0.62 (95% CI 0.60-0.64), while the trait-model still lacked predictive power, with an AUC of 0.51 (95% CI 0.49-0.54) (Figure 3E, Table 2).

The PC model did not replicate in the GenRisk study, showing no significant findings. The AUC was 0.50 (95% CI 0.47-0.53)(Table 2), with a sensitivity of 81%, specificity of 27%, and an overall accuracy of 43% (Figure 3C). Similarly, the PC case-model, including age, showed limited predictive capability with an AUC of 0.50 (95% CI 0.46-0.53). Meanwhile, the traits-model, including age, also did not demonstrate significant predictive power, with an AUC of 0.54 (95% CI 0.50 - 0.57) (Figure 3B, Table 2). Conversely, when age was excluded from the models, the AUC of both the all-model and the BC-model surprisingly improved significantly to 0.56 (95% CI 0.53 - 0.59) and 0.58 (95% CI 0.55-0.62), respectively. Meanwhile, the trait-model still lacks predictive power, with an AUC of 0.53 (95% CI 0.49-0.56) (Figure 3E, Table 2).

Considering BC and PC results, where excluding age improved the model slightly, we found that this was due to the GenRisk study design, that had a common pool of controls for all the cases, selected in an overall frequency matched design. As expected, the median ages in the UK cohort were always greater for cases than controls. However, in the GenRisk study, BC and PC cases were younger than controls, which made the age model parameter trained in the UKB reduce the predictive accuracy in the validation (Table 1. Supplementary Table 5).

## Discussion

Cancer development results from intricate interactions between genetic factors and modifiable exposures like lifestyle, diet, stress, as well as social factors such as age, sex and ancestry. An effective prevention strategy necessitates a comprehensive approach that recognizes the full spectrum of genetic and environmental influences on cancer risk. Current individual PRS often fall short in capturing this complexity, emphasizing the need for a thorough perspective to guide more effective prevention and intervention strategies.

To overcome these limitations, our study adopts a novel approach to cancer prediction, termed PRS constellation, that uses a Bayesian network model to define which PRS from diverse lifestyle and environmental factors, should be integrated and offer a more precise tool for assessing cancer risk in the context of precision prevention. This approach aimed to introduce a different method for integrating lifestyle and health-related data, especially in situations where this data may be inaccessible or when only genetic information is available, offering a distinct approach to conventional methods. Furthermore, this methodology holds promise for identifying new traits targets for prevention and diagnosis, as well as for developing personalized plans that incorporate genetic and lifestyle data.

The results from the model combining cancer PRS with risk factor PRS did not show an enhancement in predictive capability compared to the specific cancer PRS in cases of CRC and BC. While a minor enhancement was observed in PC cases, this was not been confirmed in the GenRisk study. The PC PRS revealed a limited predictive capacity in the GenRisk study, underscoring the disparities between the two populations. This suggests that further refinement is needed in defining a PRS for PC within the Spanish population. To our knowledge, no article in the literature has utilized a similar methodology for combining PRS of risk factors with PRS of cancer.

Despite these negative results, it is evident that the PRS of the selected traits collectively exhibit autonomous predictive power in the UKB, though this may be related to fact that the PRSs were developed in the UKB, and these factors indeed contribute to risk prediction. The validation in the GenRisk study failed, probably because the predictive power of the PRSs regarding the real exposures is low and this diluted effect reduced power. Furthermore, it is important to highlight the significant impact of including age and sex in the models. Specifically, the inclusion of age and sex in the colorectal cancer (CRC) model and age in the prostate cancer (PC) model resulted in an increase in the area under the curve (AUC) from 0.63 to 0.74 and from 0.60 to 0.75, respectively, in the UK Biobank (UKB) cohort. However, this trend was only observed for the CRC model in the GenRisk study, where the inclusion of age and sex only slightly improved the AUC from 0.62 to 0.65, but the AUC decreased for the BC and PC models, probably due to the frequency matched study design in the GenRisk study that lead to a wrong age distribution compared to that of the general population. For this reason, regarding assessment of the models predictive accuracy in the GenRisk study, probably the models that do not include age or sex are more appropriate.

Notably, some of the selected variables by the Bayesian network approach have not been previously identified as risk factors for the cancers studied, suggesting that our approach may uncover novel predictors. These findings highlight the potential for PRS to contribute to cancer risk assessment and warrant further investigation and validation. The final CRC PRS model incorporated 9 PRS traits, along with sex and age variables. Most selected variables and their direction of associations are consistent with previously described CRC risk factors in multiple studies, reassuring the model’s building strategy. Elevated risk of CRC has been associated with cigarette smoking, increased adult height a higher waisthip ratio (WHR), positive correlations with diabetes, hypertension, pulse rate, and elevated LDL cholesterol levels have also been identified as contributing factors(21,22). A low red blood cell count (anemia) is recognized as an early indicator of CRC(23), may also have a causal role, suggesting further exploration through Mendelian randomization studies. Aspirin and other non-steroidal anti-inflammatory drugs have been shown associated with a reduced risk of CRC(24). The protective effect may also be linked to aspirin’s documented role in preventing cardiovascular diseases, reducing blood pressure, and lowering pulse rates, all of which contribute to a reduced risk of colorectal cancer. These variables highlight common links between cardiovascular diseases and CRC, possibly mediated by chronic inflammation, metabolic dysregulation, and shared genetic predispositions. Addressing these risk factors through lifestyle changes and medical interventions could reduce both cardiovascular disease and CRC incidence, underscoring their importance in prevention and treatment strategies.

In the BC PRS constellation model, the integration of CRC PRS, along with 8 additional PRS variables and age, allowed for the establishment of bibliographic connections with this disease. Elevated waist circumference, height, and hypertension have been documented as factors associated with an increased risk of BC. Additionally, factors tied to women’s health, including uterine fibroids, age at menopause, and postmenopausal testosterone levels, exhibit positive correlations with BC risk. Conversely, age at menarche and sex hormone-binding globulin (SHBG) levels were inversely related to BC risk, as supported by relevant literature(22,25–27).

For PC, the PRS constellation model integrated 10 PRS variables (1 PRS specific to PC + 9 PRS associated with various traits), along with age. Factors such as elevated alcohol intake, increased WHR, hypertension, prostate hyperplasia, and a higher platelet cell count were positively associated with an increased risk of PC. Interestingly, although there is no evidence of a direct association between height and PC risk, its connection with higher PC mortality has been reported. On the other hand, the model retained coffee intake and whole grain consumption PRSs that were negatively associated with PC risk. High coffee intake has been previously associated with a lower risk of PC, while whole grain consumption has been linked to a lower risk of total and site-specific cancers, there is currently no evidence of its relationship with PC. Lastly, despite our results, circulating vitamin D levels have documented to be associated with better progression and prognosis of PC(22,28–32).

These findings align with current international public health guidelines highlighting that a reduced adherence to lifestyle recommendations is linked to an elevated overall cancer risk(22,33). Additionally, the results align with earlier research exploring interactions between cumulative genetic susceptibility and various variables, in the CRC(34), BC(35), and PC(36).

This study is subject to certain limitations, particularly the PRSs used to predict traits were developed in the UK Biobank, and UK Biobank samples contributed to the GWAS meta-analyses that defined the cancer specific PRSs. This may result in an overestimation of the models’ predictive accuracy. We tested this in the independent GenRisk study, suggesting challenges in translating PRS models across diverse populations, mainly for PC. To enhance the model, calculating the PRS in another Spanish cohort and subsequently validating it with GenRisk could provide more robust and generalizable results. The already mentioned issue with the age and sex matched design in GenRisk is also a relevant limitation, and ideally the PRS predictive accuracy should be tested in studies with cohort design to properly include age and sex effects.

In conclusion, the introduction of the PRS constellation model had a significant value in the UKB cohort but did not significantly enhance the predictive capabilities of cancer-specific PRS and this could not be validated in the GenRisk study. While our approach did not improve overall predictive power, it underscores the relevance of lifestyle-related factors and the necessity of incorporating these elements into cancer prevention strategies. Our study also reveals the model’s limitations, indicating the need for additional biomarkers and a more refined selection of training and validation populations to enhance efficacy. These findings emphasize the ongoing need for comprehensive approaches in advancing precision medicine and optimizing early cancer detection.

## Additional Information

## Acknowledgments

This research has been conducted using the UK Biobank Resource under Application Number 69063. We would like to thank the participants in the MCC-Spain/GenRisk project. We acknowledge the contribution of the COST Action CA21169, supported by COST (European Cooperation in Science and Technology). We want to particularly acknowledge the patients and the Biobank HUB-ICO-IDIBELL (PT17/0015/0024) for their collaboration. We thank CERCA Programme / Generalitat de Catalunya for institutional support.

## Authors’ contributions

Recruitment and acquisition of data: E.G,

Concept and design: V.M, N.M and A.V.

Preparation of genetic data: N.M and A.D.

Data curation: N.M, A.D, B.R, L.R,

Data analysis: N.M. Drafting of the manuscript: N.M.

Obtaining fundings: V.M, S.M,

Contributions to the final version of the manuscript were made by all authors.

## Ethics approval and consent to participate

UK Biobank study received ethical approval from the North West Centre for Research Ethics Committee (11/NW/0382) and had obtained written informed consent from all participants. The protocol of GenRisk was approved by each of the ethics committees of the participating institutions. The specific study reported here was approved by the Bellvitge Hospital Ethics Committee with reference PR020/17. Informed consent was obtained from all individual participants included in the study.

## Consent for publication

Not applicable.

## Data availability

This study utilizes information sourced from the UK Biobank repository (application number 69033), accessible upon request submission to the UK Biobank. For further details, visit https://www.ukbiobank.ac.uk. The GenRisk data can be obtained for research purposes applying for access to the corresponding author, subject to approval by the GenRisk Data Access Committee.

## Supplementary data

<u>Supplementary Figure 1.</u> Distribution of unweighted polygenic risk score of: A) Colorectal, B) Breast, and C) Prostate cancer, displayed for cases and controls in the UK Biobank cohort.

<u>Supplementary Figure 2.</u> AUC Analysis of PRS Constellation Prediction Models for CRC, BC, and PC (Internal Validation). ROC curves and AUC values for the prediction models.

<u>Supplementary Figure 3.</u> AUC Analysis of PRS Constellation Prediction Models for CRC, BC, and PC (Internal Validation). Individual and cumulative contributions of each factor.

<u>Supplementary Figure 4.</u> SNP correlation analysis between CRC and traits incorporated in the CRC model.

<u>Supplementary Figure 5.</u> SNP correlation analysis between BC and traits incorporated in the BC model.

<u>Supplementary Figure 6.</u> SNP correlation analysis between PC and traits incorporated in the PC model.

<u>Supplementary Table 1.</u> Collection of Traits with Calculated PRS.

<u>Supplementary Table 2.</u> Overview of the relevant information regarding genotyping, quality control, and imputation of the projects comprising the GenRisk study.

<u>Supplementary Table 3.</u> Evaluation and Selection of PRSs for Cancer and Associated Risk Traits. List of PRS calculated and type of cancer associated.

<u>Supplementary Table 4.</u> Presents predictive performance metrics for CRC, BC, and PC prediction models, including final models and 10-fold cross-validation results.

<u>Supplementary Table 5.</u> Sex and age characteristics of the GenRisk study participants.

