## Supplemental_Figures for "Multi-Cancer PRS Constellation Model for Cancer Risk Prediction"

### Supplementary Figure 1

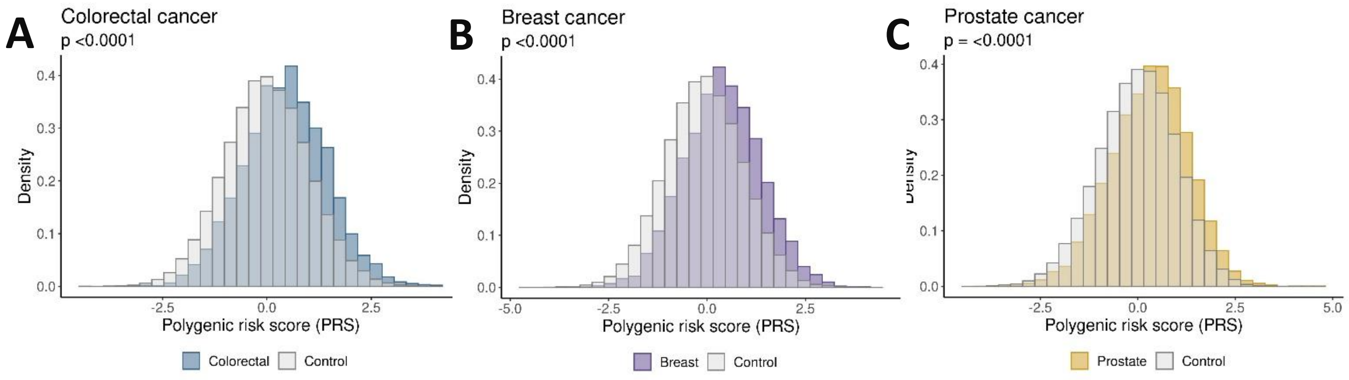


**Supplementary Figure 1.** Distribution of weighted polygenic risk score of: **A)** Colorectal, **B)** Breast, and **C)** Prostate cancer, displayed for cases and controls in the UK Biobank database.

### Supplementary Figure 2

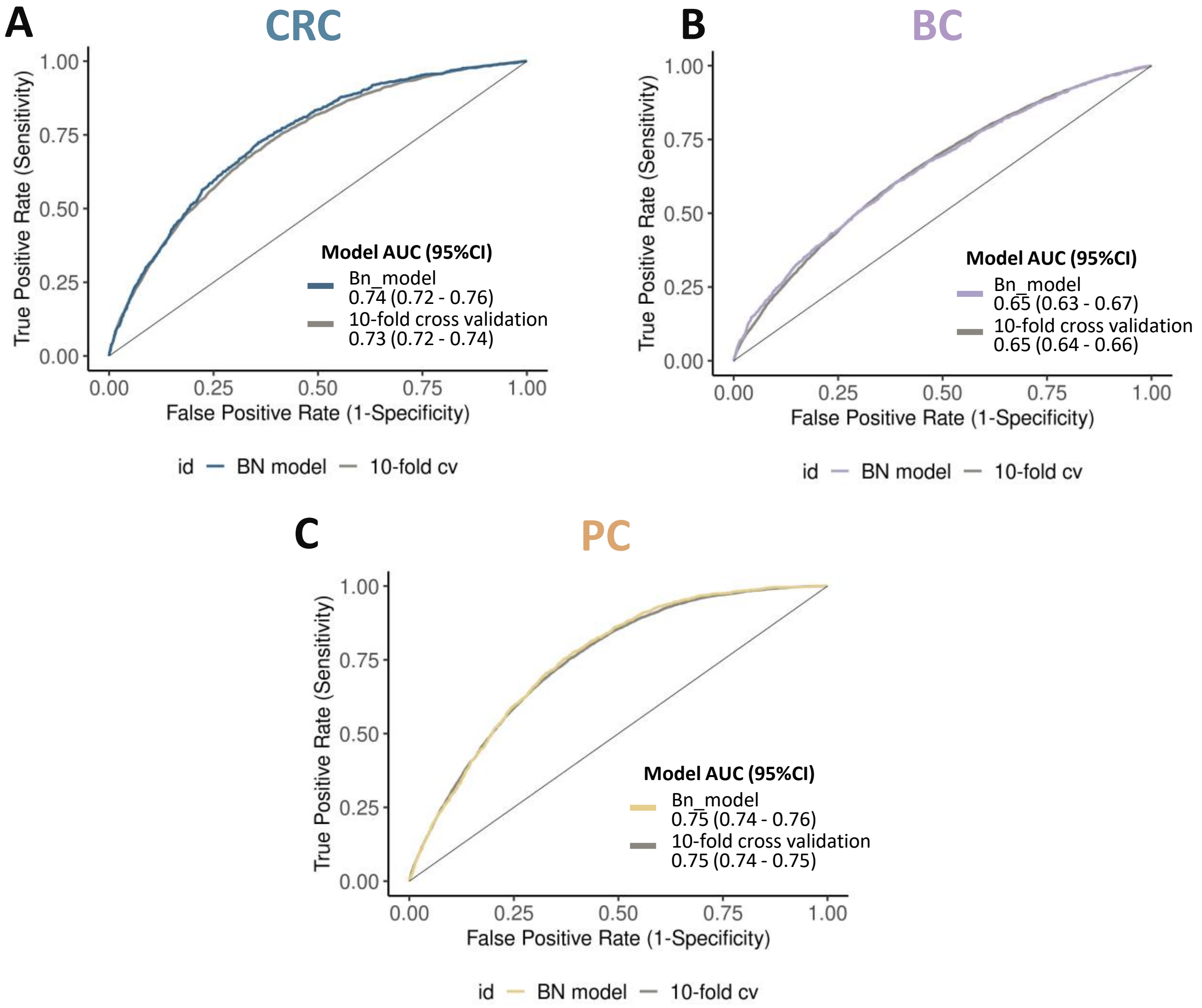


**Supplementary Figure 2. AUC Analysis of PRS Constellation Prediction Models for CRC, BC, and PC (Internal Validation)** A-C) ROC curves and AUC values for the prediction models of A) CRC, B) BC, and C) PC.

### Supplementary Figure 3

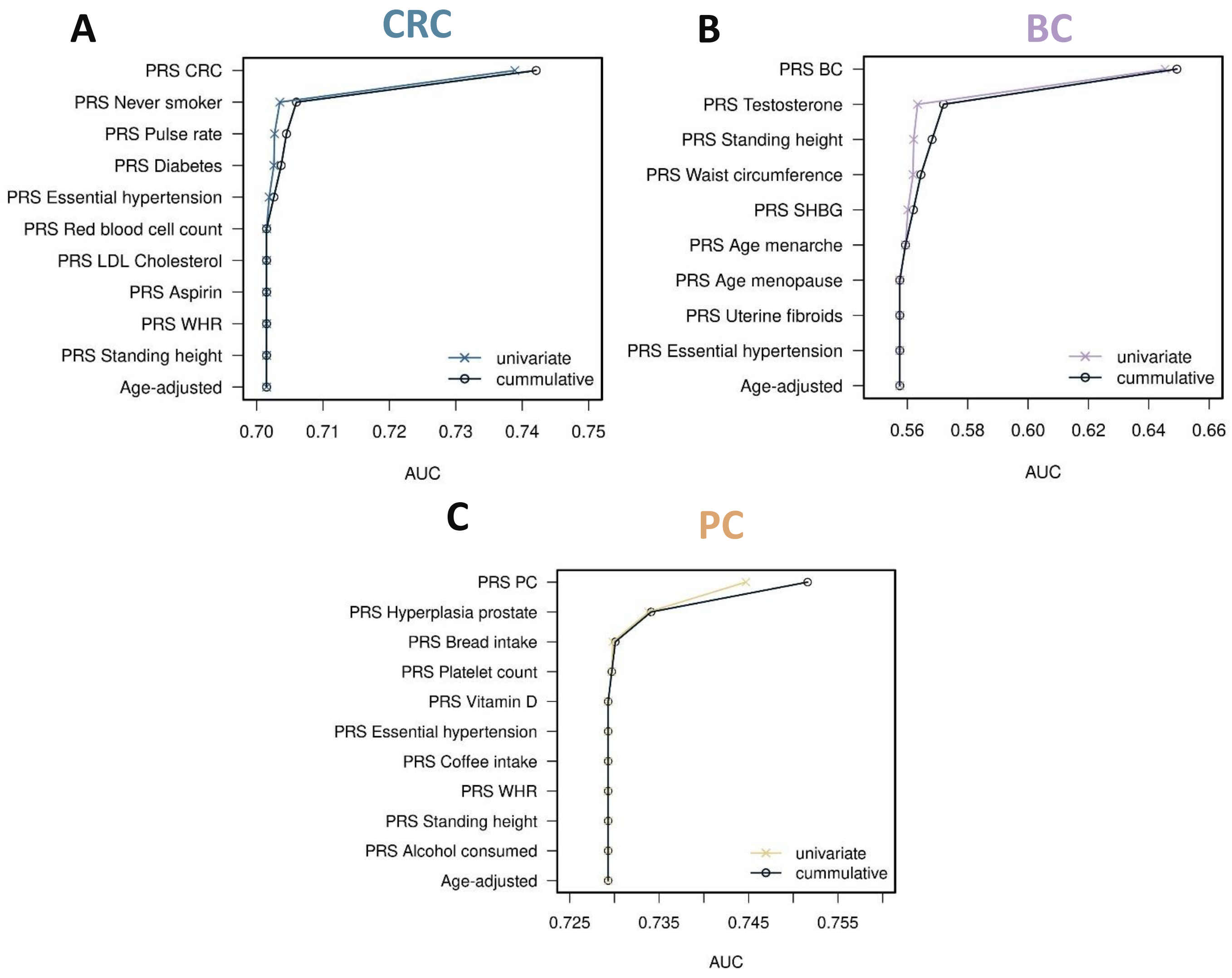


**Supplementary Figure 3. AUC Analysis of PRS Constellation Prediction Models for CRC, BC, and PC (Internal Validation).** A-C) Individual and cumulative contributions of each factor to A) CRC, B) BC, and C) PC models' predictive accuracy, represented by individual and cumulative AUCs. Cumulative AUCs are shown in gray, while individual AUCs are depicted in color. PRSs are sorted by increasing AUC. CRC = Colorectal Cancer. BC = Breast cancer, PC = Prostate cancer. WHR = Waist to hip ratio. BMI = Body mass index. BM = Biomarker. BP = Blood Pressure.

### Supplementary Figure 4

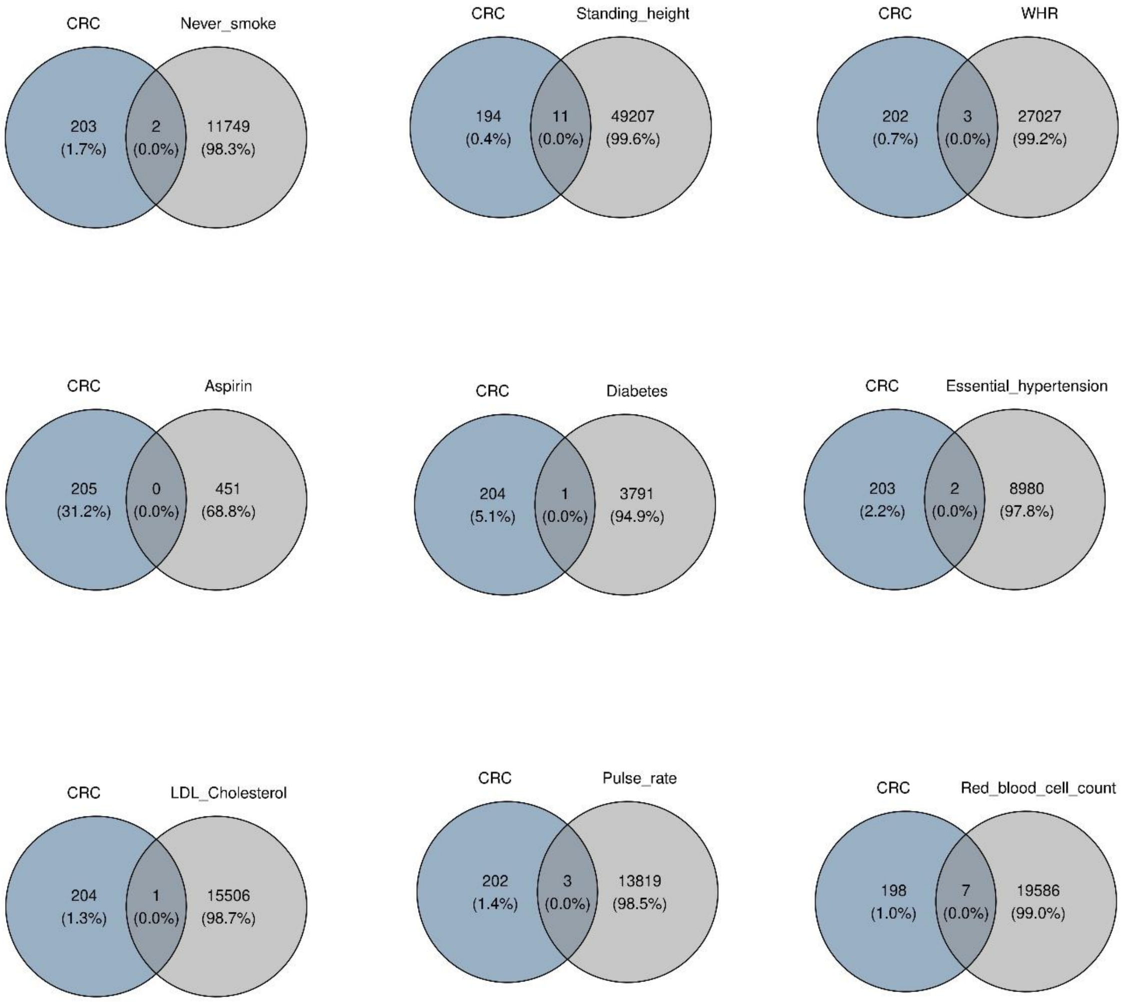


**Supplementary Figure 4. SNP correlation analysis between CRC and traits incorporated in the CRC model.** The Venn diagram illustrates the overlap of CRC-associated SNPs with the relevant CRC traits included in the model.

### Supplementary Figure 5

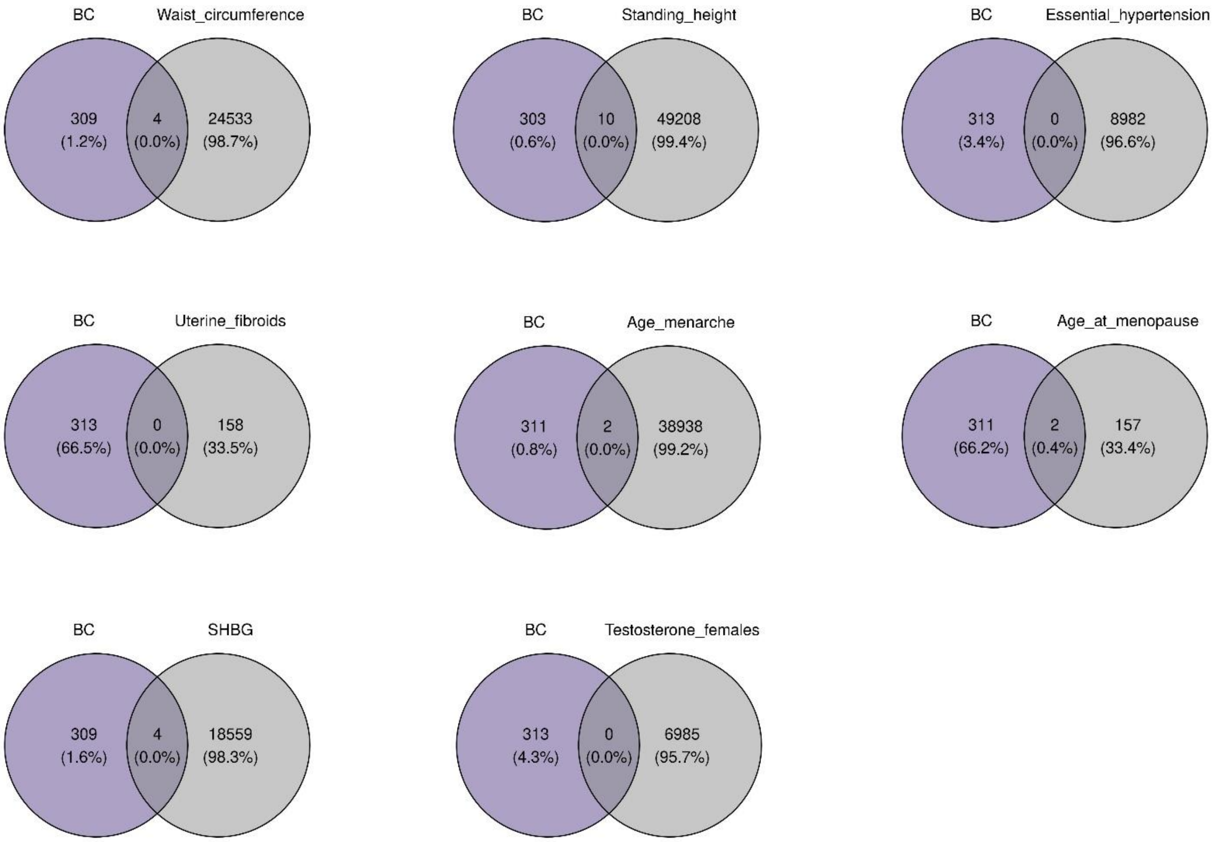


**Supplementary Figure 5. SNP correlation analysis between BC and traits incorporated in the BC model.** The Venn diagram illustrates the overlap of BC-associated SNPs with the relevant BC traits included in the model.

### Supplementary Figure 6

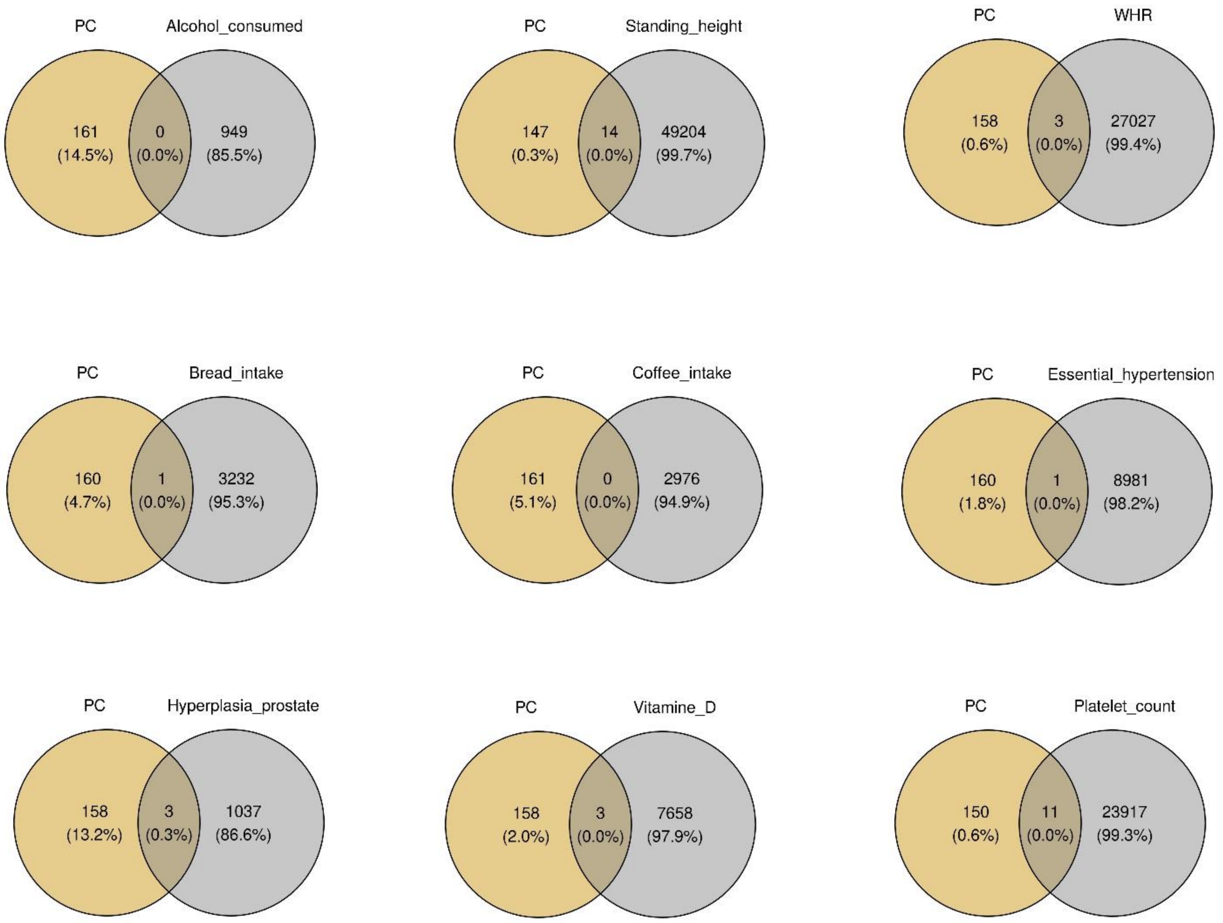


**Supplementary Figure 6. SNP correlation analysis between PC and traits incorporated in the PC model.** The Venn diagram illustrates the overlap of PC-associated SNPs with the relevant PC traits included in the model.
