## Supplemental_Tables for "Multi-Cancer PRS Constellation Model for Cancer Risk Prediction"

**Supplementary Table 1.** Collection of Traits with Calculated PRS. The following table presents information regarding the collection of traits for which PRS calculated. The columns include the trait name, trait category, the cohort of individuals in which it has been calculated (Cohort), the number of SNPs included (#Variants in PRS), phenotype ID if part of the PGS catalog(PGS Catalog ID), the ethnic category of this population, and for which of the three types of cancer studied the trait is a risk factor (Risk factor of). CRC: Colorectal cancer; BC: Breast cancer; PC: Prostate cancer; FH: Family history; AR - automated reading; UKB: UK Biobank; GECCO: German Corona Consensus Dataset; CORECT: Colorectal Transdisciplinary Study.

| # | Trait Name | Trait Category | Nº Variants in PRS | PGS Catalog ID | GWAS Ethnicity | Risk factor of | First author and year | Link to publication |
|---|---|---|---|---|---|---|---|---|
| 1 | Colorectal cancer | Cancer | 204 | - | European ancestry | CRC | Ceres Fernandez-Rozadilla *et al.* | https://www.nature.com/articles/s41588-022-01222-9#MOESM1 |
| 2 | Breast cancer | Cancer | 313 | - | European ancestry | BC | Nasim Mavaddat *et al.* | https://www.cell.com/ajhg/fulltext/S0002-9297(18)30405-1 |
| 3 | Prostate cancer | Cancer | 161 | PGS000084 | European: 93.3%, African: | PC | Rebecca E. Graff *et al.* | https://www.nature.com/articles/s41467-021-21288-z |
| 4 | Alcohol intake frequency | Alcohol consumption measurement | 8353 | PGS001087 | European ancestry | CRC/BC/PC | Yosuke Tanigawa *et al.* | https://journals.plos.org/plosgenetics/article?id=10.1371/journal.pgen.1010105 |
| 5 | Alcohol consumed | Alcohol consumption measurement | 1028 | PGS001086 | European ancestry | CRC/BC/PC | Yosuke Tanigawa *et al.* | https://journals.plos.org/plosgenetics/article?id=10.1371/journal.pgen.1010105 |
| 6 | Freq. of drinking alcohol | Alcohol consumption measurement | 860 | PGS001394 | European ancestry | CRC/BC/PC | Yosuke Tanigawa *et al.* | https://journals.plos.org/plosgenetics/article?id=10.1371/journal.pgen.1010105 |
| 7 | Average weekly alcohol consumption (beer and cider) | Alcohol consumption measurement | 2842 | PGS001088 | European ancestry | CRC/BC/PC | Yosuke Tanigawa *et al.* | https://journals.plos.org/plosgenetics/article?id=10.1371/journal.pgen.1010105 |
| 8 | Smoking status | Smoking consumption measurement | 974 | PGS001129 | European ancestry | CRC/BC/PC | Yosuke Tanigawa *et al.* | https://journals.plos.org/plosgenetics/article?id=10.1371/journal.pgen.1010105 |
| 9 | Number of cigarettes previously smoked daily (current cigar/pipe smokers) | Smoking consumption measurement | 725 | PGS001130 | European ancestry | CRC/BC/PC | Yosuke Tanigawa *et al.* | https://journals.plos.org/plosgenetics/article?id=10.1371/journal.pgen.1010105 |
| 10 | Never Smoker | Smoking consumption measurement | 12310 | PGS001127 | European ancestry | CRC/BC/PC | Yosuke Tanigawa *et al.* | https://journals.plos.org/plosgenetics/article?id=10.1371/journal.pgen.1010105 |
| 11 | Weight | Body Mesurements | 31222 | PGS001230 | European ancestry | CRC/BC/PC | Yosuke Tanigawa *et al.* | https://journals.plos.org/plosgenetics/article?id=10.1371/journal.pgen.1010105 |
| 12 | BMI (body mass index) | Body Mesurements | 27126 | PGS001228 | European ancestry | CRC/BC/PC | Yosuke Tanigawa *et al.* | https://journals.plos.org/plosgenetics/article?id=10.1371/journal.pgen.1010105 |
| 13 | Waist circumference | Body Mesurements | 25538 | PGS001227 | European ancestry | CRC/BC/PC | Yosuke Tanigawa *et al.* | https://journals.plos.org/plosgenetics/article?id=10.1371/journal.pgen.1010105 |
| 14 | Hip circumference | Body Mesurements | 24605 | PGS001162 | European ancestry | CRC/BC/PC | Yosuke Tanigawa *et al.* | https://journals.plos.org/plosgenetics/article?id=10.1371/journal.pgen.1010105 |
| 15 | Standing height | Body Mesurements | 51209 | PGS001229 | European ancestry | CRC/BC/PC | Yosuke Tanigawa *et al.* | https://journals.plos.org/plosgenetics/article?id=10.1371/journal.pgen.1010105 |
| 16 | Body fat % | Body Mesurements | 27396 | PGS001101 | European ancestry | CRC/BC/PC | Yosuke Tanigawa *et al.* | https://journals.plos.org/plosgenetics/article?id=10.1371/journal.pgen.1010105 |
| 17 | Waist-hip ratio | Body Mesurements | 27033 | PGS002477 | European: 25%/ African: 2 | CRC/BC/PC | Omer Weissbrod *et al.* | https://www.nature.com/articles/s41588-022-01036-9 |
| 18 | Number of days/week walked 10+ minutes | Physical Activity | 400 | Gene Atlas | European ancestry | CRC/BC/PC | Oriol Canela-Xandri *et al.* | https://www.nature.com/articles/s41588-018-0248-z |
| 19 | Duration of Walks | Physical Activity | 1323 | PGS001073 | European ancestry | CRC/BC/PC | Yosuke Tanigawa *et al.* | https://journals.plos.org/plosgenetics/article?id=10.1371/journal.pgen.1010105 |
| 20 | Fresh fruit intake | Diet measurament | 4257 | PGS001062 | European ancestry | CRC/BC/PC | Yosuke Tanigawa *et al.* | https://journals.plos.org/plosgenetics/article?id=10.1371/journal.pgen.1010105 |
| 21 | Cooked vegetable intake | Diet measurament | 1085 | PGS001061 | European ancestry | CRC/BC/PC | Yosuke Tanigawa *et al.* | https://journals.plos.org/plosgenetics/article?id=10.1371/journal.pgen.1010105 |
| 22 | Oily fish intake | Diet measurament | 3925 | PGS000993 | European ancestry | CRC/BC/PC | Yosuke Tanigawa *et al.* | https://journals.plos.org/plosgenetics/article?id=10.1371/journal.pgen.1010105 |
| 23 | Processed meat intake | Diet measurament | 2346 | PGS001067 | European ancestry | CRC/BC/PC | Yosuke Tanigawa *et al.* | https://journals.plos.org/plosgenetics/article?id=10.1371/journal.pgen.1010105 |
| 24 | Beef intake | Diet measurament | 991 | PGS001056 | European ancestry | CRC/BC/PC | Yosuke Tanigawa *et al.* | https://journals.plos.org/plosgenetics/article?id=10.1371/journal.pgen.1010105 |
| 25 | Poultry intake | Diet measurament | 1609 | PGS001066 | European ancestry | CRC/BC/PC | Yosuke Tanigawa *et al.* | https://journals.plos.org/plosgenetics/article?id=10.1371/journal.pgen.1010105 |
| 26 | Milk type: Skimmed | Diet measurament | 116 | PGS001064 | European ancestry | CRC/BC/PC | Yosuke Tanigawa *et al.* | https://journals.plos.org/plosgenetics/article?id=10.1371/journal.pgen.1010105 |
| 27 | Bread intake | Diet measurament | 3483 | PGS000978 | European ancestry | CRC/BC/PC | Yosuke Tanigawa *et al.* | https://journals.plos.org/plosgenetics/article?id=10.1371/journal.pgen.1010105 |
| 28 | Cereal intake | Diet measurament | 4882 | PGS001057 | European ancestry | CRC/BC/PC | Yosuke Tanigawa *et al.* | https://journals.plos.org/plosgenetics/article?id=10.1371/journal.pgen.1010105 |
| 29 | Coffee intake | Diet measurament | 3154 | PGS001126 | European ancestry | CRC/BC/PC | Yosuke Tanigawa *et al.* | https://journals.plos.org/plosgenetics/article?id=10.1371/journal.pgen.1010105 |
| 30 | Coffee consumed | Diet measurament | 48 | PGS001123 | European ancestry | CRC/BC/PC | Yosuke Tanigawa *et al.* | https://journals.plos.org/plosgenetics/article?id=10.1371/journal.pgen.1010105 |
| 31 | Glucosamine | Supplement medicaments exposure | 324 | PGS001044 | European ancestry | CRC/BC/PC | Yosuke Tanigawa *et al.* | https://journals.plos.org/plosgenetics/article?id=10.1371/journal.pgen.1010105 |
| 32 | Taking other prescription medications | Supplement medicaments exposure | 1846 | PGS001118 | European ancestry | CRC/BC/PC | Yosuke Tanigawa *et al.* | https://journals.plos.org/plosgenetics/article?id=10.1371/journal.pgen.1010105 |
| 33 | Nº of medications taken (Number of treatments/medications taken) | Supplement medicaments exposure | 8085 | PGS001003 | European ancestry | CRC/BC/PC | Yosuke Tanigawa *et al.* | https://journals.plos.org/plosgenetics/article?id=10.1371/journal.pgen.1010105 |
| 34 | Paracetamol (Treatment/medication code) | Supplement medicaments exposure | 4673 | PGS001115 | European ancestry | CRC/BC/PC | Yosuke Tanigawa *et al.* | https://journals.plos.org/plosgenetics/article?id=10.1371/journal.pgen.1010105 |
| 35 | Ibuprofen (e.g. Nurofen) | Supplement medicaments exposure | 174 | PGS001116 | European ancestry | CRC/BC/PC | Yosuke Tanigawa *et al.* | https://journals.plos.org/plosgenetics/article?id=10.1371/journal.pgen.1010105 |
| 36 | Aspirin | Supplement medicaments exposure | 499 | PGS001112 | European ancestry | CRC/BC/PC | Yosuke Tanigawa *et al.* | https://journals.plos.org/plosgenetics/article?id=10.1371/journal.pgen.1010105 |
| 37 | Diabetes (Diabetes diagnosis ) | Medical history | 4053 | PGS001327 | European ancestry | CRC/BC/PC | Yosuke Tanigawa *et al.* | https://journals.plos.org/plosgenetics/article?id=10.1371/journal.pgen.1010105 |
| 38 | Age diabetes diagnosed | Medical history | 26 | PGS001371 | European ancestry | CRC/BC/PC | Yosuke Tanigawa *et al.* | https://journals.plos.org/plosgenetics/article?id=10.1371/journal.pgen.1010105 |
| 39 | Essential hypertension | Medical history | 9400 | PGS000958 | European ancestry | CRC/BC/PC | Yosuke Tanigawa *et al.* | https://journals.plos.org/plosgenetics/article?id=10.1371/journal.pgen.1010105 |
| 40 | Cholesterol (biomarker) | Medical history | 17204 | PGS000677 | European ancestry | CRC/BC/PC | Nasa Sinnott-Armstrong *et al.* | https://www.nature.com/articles/s41588-020-00757-z |
| 41 | HDL cholesterol (biomarker) | Medical history | 25069 | PGS000686 | European ancestry | CRC/BC/PC | Nasa Sinnott-Armstrong *et al.* | https://www.nature.com/articles/s41588-020-00757-z |
| 42 | LDL cholesterol (biomarker) | Medical history | 16184 | PGS000688 | European ancestry | CRC/BC/PC | Nasa Sinnott-Armstrong *et al.* | https://www.nature.com/articles/s41588-020-00757-z |
| 43 | Crohns disease (self-reported) | Medical history | 257 | PGS001331 | European ancestry | CRC | Yosuke Tanigawa *et al.* | https://journals.plos.org/plosgenetics/article?id=10.1371/journal.pgen.1010105 |
| 44 | Ulcerative colitis (self-reported) | Medical history | 179 | PGS001306 | European ancestry | CRC | Yosuke Tanigawa *et al.* | https://journals.plos.org/plosgenetics/article?id=10.1371/journal.pgen.1010105 |
| 45 | Diverticular disease/diverticulitis (self-reported) | Medical history | 368 | PGS000996 | European ancestry | CRC | Yosuke Tanigawa *et al.* | https://journals.plos.org/plosgenetics/article?id=10.1371/journal.pgen.1010105 |
| 46 | Malabsorption/coeliac disease (self-reported) | Medical history | 428 | PGS001301 | European ancestry | CRC | Yosuke Tanigawa *et al.* | https://journals.plos.org/plosgenetics/article?id=10.1371/journal.pgen.1010105 |
| 47 | Pulse rate (AR) | Medical history | 14455 | PGS001233 | European ancestry | CRC/BC/PC | Yosuke Tanigawa *et al.* | https://journals.plos.org/plosgenetics/article?id=10.1371/journal.pgen.1010105 |
| 48 | Uterine fibroids | Women's / Men health | 161 | PGS001032 | European ancestry | BC | Yosuke Tanigawa *et al.* | https://journals.plos.org/plosgenetics/article?id=10.1371/journal.pgen.1010105 |
| 49 | Age when periods started (menarche) | Women's / Men health | 38940 | PGS001915 | European ancestry | BC | Florian Privé *et al.* | https://www.cell.com/ajhg/fulltext/S0002-9297(21)00420-1 |
| 50 | Age at menopause (last menstrual period) | Women's / Men health | 159 | PGS002821 | European ancestry | BC | Ying Ma *et al.* | https://www.cell.com/ajhg/fulltext/S0002-9297(22)00404-9 |
| 51 | Length of menstrual cycle | Women's / Men health | 339 | PGS001913 | European ancestry | BC | Florian Privé *et al.* | https://www.cell.com/ajhg/fulltext/S0002-9297(21)00420-1 |
| 52 | Number of live births | Women's / Men health | 1639 | PGS002381 | European: 25%/ African: 2 | BC | Omer Weissbrod *et al.* | https://www.nature.com/articles/s41588-022-01036-9 |
| 53 | Hyperplasia of prostate | Women's / Men health | 1223 | PGS001338 | European ancestry | PC | Yosuke Tanigawa *et al.* | https://journals.plos.org/plosgenetics/article?id=10.1371/journal.pgen.1010105 |
| 54 | Enlarged prostate | Women's / Men health | 449 | PGS001015 | European ancestry | PC | Yosuke Tanigawa *et al.* | https://journals.plos.org/plosgenetics/article?id=10.1371/journal.pgen.1010105 |
| 55 | Vitamin D (nmol/L) | Biomarkers | 8012 | PGS000702 | European ancestry | CRC/BC/PC | Nasa Sinnott-Armstrong *et al.* | https://www.nature.com/articles/s41588-020-00757-z |
| 56 | Glucose(mmol/L) | Biomarkers | 3313 | PGS000684 | European ancestry | CRC/BC/PC | Nasa Sinnott-Armstrong *et al.* | https://www.nature.com/articles/s41588-020-00757-z |
| 57 | Red blood cell count (10^12 cells/Litre.) | Biomarkers | 20480 | PGS001240 | European ancestry | CRC/BC/PC | Yosuke Tanigawa *et al.* | https://journals.plos.org/plosgenetics/article?id=10.1371/journal.pgen.1010105 |
| 58 | Platelet count (10^9 cells/Litre) | Biomarkers | 24893 | PGS001238 | European ancestry | CRC/BC/PC | Yosuke Tanigawa *et al.* | https://journals.plos.org/plosgenetics/article?id=10.1371/journal.pgen.1010105 |
| 59 | Lymphocyte count | Biomarkers | 4212 | PGS001199 | European ancestry | CRC/BC/PC | Yosuke Tanigawa *et al.* | https://journals.plos.org/plosgenetics/article?id=10.1371/journal.pgen.1010105 |
| 60 | Monocyte count | Biomarkers | 9323 | PGS001163 | European ancestry | CRC/BC/PC | Yosuke Tanigawa *et al.* | https://journals.plos.org/plosgenetics/article?id=10.1371/journal.pgen.1010105 |
| 61 | Globulina de transport de les hormones sexuals (variable "SHBG"). | Biomarkers | 19328 | PGS000694 | European ancestry | BC | Nasa Sinnott-Armstrong *et al.* | https://www.nature.com/articles/s41588-020-00757-z |
| 62 | Testosterone (Only in females) | Biomarkers | 7168 | PGS000322 | White British individuals | BC | Emily Flynn *et al.* | https://www.nature.com/articles/s41431-020-00712-w |
| 63 | Testosterone (male only) | Biomarkers | 3985 | PGS001988 | European ancestry | PC | Florian Privé *et al.* | https://www.cell.com/ajhg/fulltext/S0002-9297(21)00420-1 |

**Supplementary Table 2.** Overview of the relevant information regarding genotyping, quality control, and imputation of the projects comprising the GenRisk dataset. The following columns are included: Project name: The name of the project. Samples: The number of samples used in the project. Genotype date: The date of genotyping. Array: The genotyping array used. SNPs included: The number of SNP included in teh array. SNPs-QC: The number of SNP after quality control (QC). MCC-Spain samples: the number of samples belonging to MCC spain. Control: The number of control samples. Case: The number of case samples. First author: The primary author of the reference article. Link to publication: A link to the publication related to the project.

| Project name: | Samples | | | | | | MCC-Spain samples | Genotipe date: | Array: | SNPs markers included | SNPs-QC | Project name: | First author | Link to publication |
|---|---|---|---|---|---|---|---|---|---|---|---|---|---|---|
| | **Breast** | **CLL** | **Colorectal** | **Control** | **Gastric** | **Prostate** | | | | | | | | |
| CRCGen | 0 | 0 | 541 | 485 | 0 | 0 | 1026 | 2016 | Infinium Oncoarray-500 k | 533631 | 451568 | CRCGen | Mireia Obón-Santacana *et al.* | https://www.ncbi.nlm.nih.gov/pmc/articles/PMC8574048/ |
| GASTRIC | 0 | 0 | 0 | 0 | 225 | 0 | 225 | 2016 | Illumina Infinium Omni2,5 Exome v1,3 | 2612357 | 2592364 | CNIO | Stephanie L Schmit et al. | https://academic.oup.com/jnci/article/111/2/146/5039592?login=false |
| PRACTICAL | 0 | 0 | 0 | 402 | 0 | 527 | 929 | 2018 | Infinium Oncoarray-500 k | 533631 | 485120 | PRACTICAL | Schumacher FR et al. | https://www.nature.com/articles/s41588-018-0142-8 |
| CNIO | 1179 | 390 | 890 | 1977 | 101 | 386 | 4923 | 2020/2021 | Illumina Global Screening Array-24 v3.0 | 660945 | 657992 | CNIO/CRCGen/PRACTICAL | National Cancer Institute | https://epi.grants.cancer.gov/gameon/ |
| | | | | | | | **7103** | | | | | | | |

**Supplementary Table 3. Evaluation and Selection of PRSs for Cancer and Associated Risk Traits.** List of PRS calculated and type of cancer associated. PRS evaluation: Assessment of PRS predictive power for corresponding trait.

| | | | **PRS evaluation** | |
|---|---|---|---|---|
| **#** | **Trait Name** | **Risk factor of** | **Test** | **p.value** |
| 1 | Colorectal cancer | **CRC** | t.test | 8.0E-160 |
| 2 | Breast cancer | **BC** | t.test | 1.8E-226 |
| 3 | Prostate cancer | **PC** | t.test | <0.0001 |
| 4 | Alcohol intake frequency | **CRC/BC/PC** | ANOVA | <0.0001 |
| 5 | Alcohol consumed | **CRC/BC/PC** | t.test | 1.1E-239 |
| 6 | Freq. of drinking alcohol | **CRC/BC/PC** | ANOVA | <0.0001 |
| 7 | Average weekly alcohol consumption (beer and cider) | **CRC/BC/PC** | ANOVA | <0.0001 |
| 8 | Smoking status | **CRC/BC/PC** | ANOVA | <0.0001 |
| 9 | Number of cigarettes previously smoked daily (current cigar/pipe smokers) | **CRC/BC/PC** | ANOVA | <0.0001 |
| 10 | Never Smoker | **CRC/BC/PC** | ANOVA | <0.0001 |
| 11 | Weight | **CRC/BC/PC** | Pearson | <0.0001 |
| 12 | BMI (body mass index) | **CRC/BC/PC** | Pearson | <0.0001 |
| 13 | Waist circumference | **CRC/BC/PC** | Pearson | <0.0001 |
| 14 | Hip circumference | **CRC/BC/PC** | Pearson | <0.0001 |
| 15 | Standing height | **CRC/BC/PC** | Pearson | <0.0001 |
| 16 | Body fat % | **CRC/BC/PC** | Pearson | <0.0001 |
| 17 | Waist-hip ratio | **CRC/BC/PC** | Pearson | <0.0001 |
| 18 | Number of days/week walked 10+ minutes | **CRC/BC/PC** | Pearson | 4.5E-13 |
| 19 | Duration of Walks | **CRC/BC/PC** | Pearson | <0.0001 |
| 20 | Fresh fruit intake | **CRC/BC/PC** | Pearson | <0.0001 |
| 21 | Cooked vegetable intake | **CRC/BC/PC** | Pearson | <0.0001 |
| 22 | Oily fish intake | **CRC/BC/PC** | Pearson | <0.0001 |
| 23 | Processed meat intake | **CRC/BC/PC** | Pearson | <0.0001 |
| 24 | Beef intake | **CRC/BC/PC** | Pearson | 2.9E-66 |
| 25 | Poultry intake | **CRC/BC/PC** | Pearson | <0.0001 |

| | | | | |
|---|---|---|---|---|
| 26 | Milk type: Skimmed | CRC/BC/PC | Pearson | 6.1E-50 |
| 27 | Bread intake | CRC/BC/PC | Pearson | <0.0001 |
| 28 | Cereal intake | CRC/BC/PC | Pearson | <0.0001 |
| 29 | Coffee intake | CRC/BC/PC | Pearson | 1.4E-08 |
| 30 | Coffee consumed | CRC/BC/PC | t.test | 7.6E-23 |
| 31 | Glucosamine | CRC/BC/PC | ANOVA | <0.0001 |
| 32 | Taking other prescription medications | CRC/BC/PC | t.test | <0.0001 |
| 33 | Nº of medications taken (Number of treatments/medications taken) | CRC/BC/PC | Pearson | <0.0001 |
| 34 | Paracetamol (Treatment/medication code) | CRC/BC/PC | t.test | 2.55E-257 |
| 35 | Ibuprofen (e.g. Nurofen) | CRC/BC/PC | t.test | 2.75E-73 |
| 36 | Aspirin | CRC/BC/PC | t.test | <0.0001 |
| 37 | Diabetes (Diabetes diagnosis ) | CRC/BC/PC | t.test | <0.0001 |
| 38 | Age diabetes diagnosed | CRC/BC/PC | Pearson | 1.2E-57 |
| 39 | Essential hypertension | CRC/BC/PC | Pearson | 3.9E-49 |
| 40 | Cholesterol (biomarker) | CRC/BC/PC | Pearson | <0.0001 |
| 41 | HDL cholesterol (biomarker) | CRC/BC/PC | Pearson | <0.0001 |
| 42 | LDL cholesterol (biomarker) | CRC/BC/PC | Pearson | <0.0001 |
| 43 | Crohns disease (self-reported) | CRC | t.test | 2.5E-34 |
| 44 | Ulcerative colitis (self-reported) | CRC | t.test | 1.9E-51 |
| 45 | Diverticular disease/diverticulitis (self-reported) | CRC | t.test | 7.0E-28 |
| 46 | Malabsorption/coeliac disease (self-reported) | CRC | t.test | 3.4E-80 |
| 47 | Pulse rate (AR) | CRC/BC/PC | Pearson | <0.0001 |
| 48 | Uterine fibroids | BC | t.test | 1.8E-22 |
| 49 | Age when periods started (menarche) | BC | Pearson | <0.0001 |
| 50 | Age at menopause (last menstrual period) | BC | Pearson | <0.0001 |
| 51 | Length of menstrual cycle | BC | Pearson | 2.0E-16 |
| 52 | Number of live births | BC | Pearson | <0.0001 |
| 53 | Hyperplasia of prostate | PC | t.test | 1.5E-17 |
| 54 | Enlarged prostate | PC | t.test | 1.0E-38 |
| 55 | Vitamin D (nmol/L) | CRC/BC/PC | Pearson | <0.0001 |
| 56 | Glucose(mmol/L) | CRC/BC/PC | Pearson | <0.0001 |
| 57 | Red blood cell count (10^12 cells/Litre.) | CRC/BC/PC | Pearson | <0.0001 |
| 58 | Platelet count (10^9 cells/Litre) | CRC/BC/PC | Pearson | <0.0001 |

| | | | | |
|---|---|---|---|---|
| 59 | Lymphocyte count | **CRC/BC/PC** | Pearson | <0.0001 |
| 60 | Monocyte count | **CRC/BC/PC** | Pearson | <0.0001 |
| 61 | globulina de transport de les hormones sexuals (variable "SHBG"). | **BC** | Pearson | <0.0001 |
| 62 | Testosterone (Only in females) | **BC** | Pearson | <0.0001 |
| 63 | Testosterone (male only) | **PC** | Pearson | <0.0001 |

**Supplementary Table 4.** Presents predictive performance metrics for CRC, BC, and PC prediction models, including final models and 10-fold cross-validation results.

| PRS model | Cohort | AUC | 95% CI | p.value* |
|---|---|---|---|---|
| CRC | bn_model | 0.74 | 0.72 - 0.76 | |
| | cv_f10 | 0.73 | 0.72 - 0.74 | 0.27 |
| BC | bn_model | 0.65 | 0.63 - 0.67 | |
| | cv_f10 | 0.65 | 0.64 - 0.66 | 0.95 |
| PC | bn_model | 0.77 | 0.76 - 0.79 | |
| | cv_f10 | 0.77 | 0.76 - 0.77 | 0.52 |

* p.value bn_model vs cross validations

**Supplementary Table 5. Sex and age characteristics of the GenRisk study participants.** CRC: Colorectal cancer; BC: Breast cancer; PC: Prostate cancer

| | | CRC | | BC | | PC | |
|---|---|---|---|---|---|---|---|
| | | **Control (%)** | **Case (%)** | **Control (%)** | **Case (%)** | **Control (%)** | **Case (%)** |
| **Total** | | 1977 | 890 | 1334 | 1179 | 862 | 386 |
| | | 68.96 | 31.04 | 53.1 | 46.9 | 69.1 | 30.9 |
| **Sex** | Female | 56.5 | 36.9 | 100 | 100 | | |
| | Male | 43.5 | 63.1 | | | 100 | 100 |
| **Age (years)** | ≤45 | 12.3 | 3.4 | 18.5 | 20.3 | 4.1 | 0.3 |
| | (45, 55] | 18.6 | 13.8 | 23.9 | 30.3 | 11.5 | 9.1 |
| | (55,65] | 26.2 | 28.5 | 21.7 | 26.6 | 31.0 | 42.7 |
| | >65 | 42.9 | 54.3 | 35.9 | 22.8 | 53.5 | 47.9 |
| | Median | 64 | 67 | 59 | 55 | 66 | 65 |
